# CONCEPTUAL ADVANCEMENT: SOCIAL HEALTH AS A FACILITATOR IN THE USE OF COGNITIVE RESERVE

**DOI:** 10.1101/2022.06.07.22276079

**Authors:** Myrra Vernooij-Dassen, Eline Verspoor, Suraj Samtani, Perminder Sachdev, M. Arfan Ikram, Meike W. Vernooij, Claudia Hubers, Rabih Chattat, Marta Lenart, Joanna Rymaszewska, Dorota Szczesnia, Henry Brodaty, Anna-Karin Welmer, Jane Maddock, Isabelle van der Velpen, Henrik Wiegelmann, Anna Marseglia, Marcus Richards, Rene Melis, Marjolein de Vugt, Esme Moniz-Cook, Yun-Hee Jeon, Marieke Perry, Karin Wolf-Ostermann

**Affiliations:** Scientific Center for Quality of Healthcare, Radboud University Medical Center, Nijmegen, the Netherlands; Department Geriatric Medicine, Radboud University Medical Center, Nijmegen, the Netherlands; Centre for Healthy Brain Ageing (CHeBA), Sydney, Australia; Department of Epidemiology, Erasmus University Medical Center; Harvard T.H. Chan School of Public Health (US); Department of Radiology and Nuclear Medicine, Erasmus University Medical Center; Radboud University Nijmegen, the Netherlands; University Bologna, Bologna, Italy; Department of Psychiatry, Wroclaw, Poland; Dementia Centre for Research Collaboration, Sydney, Australia; Aging Research Center, Department of Neurobiology, Care Sciences and Society, Karolinska Institutet and Stockholm University, Stockholm, Sweden; Division of Physiotherapy, Department of Neurobiology, Care Sciences and Society, Karolinska Institutet, Stockholm, Sweden; Karolinska University Hospital, Stockholm, Sweden; Stockholm Gerontology Research Center, Stockholm, Sweden; MRC Unit for Lifelong Health and Ageing at UCL, Faculty of Population Health University College London, London, UK; University Bremen, Bremen, Germany; Division of Clinical Geriatrics, Center for Alzheimer Research, Department of Neurobiology, Care Sciences and Society, Karolinska Institutet, Stockholm, Sweden; University Maastricht, Maastricht, the Netherlands; University Hull, Hull, UK; University of Sydney, Sydney, Australia

**Author notes:** Prof. Dr. Myrra Vernooij-Dassen, Radboud University Medical Center, Scientific Center for Quality of Healthcare, IQ Healthcare 114, PO Box 9101, 6500HB Nijmegen, The Netherlands, T+31611079653.

## Abstract

Dementia is a syndrome where the origins are not fully understood, and we have no cure. New thinking through exploration of paradigms beyond biological approaches has scope to improve knowledge about this complex condition. We aim to explore the role of social health in cognitive decline and the onset of dementia. We performed a scoping and a systematic review of the literature, hypothesizing that social health acts as a driver for stimulating the use of cognitive reserve. The review yielded theoretical pathways and evidence for the association between neurobiological and social health markers. However, lack of conceptual clarity on social health hinders its articulation and associated inclusion of social health markers in epidemiological studies. We therefore apply concept advancement of social health. We developed a conceptual framework, and we present a first testing of our overarching hypothesis. This framework proved to effectively guide the identification of social health markers in our epidemiological data bases. This promotes the identification of modifiable risk factors, which may in turn shape new avenues for preventive interventions.

## INTRODUCTION

New thinking beyond fixed borders is required to understand the origins of dementia since its mechanisms are unclear and there still is no cure. The recognition of dementia, including Alzheimer’s Disease, as a multifactorial disorder determined by the interplay of genetic susceptibility and environmental factors across the lifespan (Winblad et al., 2016), encourages the exploration of new pathways and hypotheses. We aim to explore the role of social health in cognitive decline and the onset of dementia. Social health has been indicated by the WHO as the social domain of health, alongside mental and physical health (WHO, 1946). According to Huber et al. social health has been conceptualised as the influence of social resources in finding a balance between capacities and limitations (Huber et al., 2011).

Our current understanding of risk for Alzheimer’s Disease is primarily driven by the amyloid cascade hypothesis (Hardy & Selkoe, 2002). While medications are able to remove amyloid from the brain, no effects on clinical outcomes have been reported (Richard, den Brok, & van Gool, 2021). This suggests that the amyloid hypothesis does not tell us the complete developmental story. The discordance found between neuropathology and clinical symptoms means that people with neuropathology do not always have clinical symptoms and that clinical symptoms do not always match with neuropathology (Winblad et al., 2016). Also acknowledgement that dementia is not ‘normal ageing’ nor is it an acceleration of ageing (Marseglia et al., 2020) suggests that neither amyloids nor ageing alone can explain the development of Alzheimer’s Disease. The latter is illustrated by new findings noting that age-specific incidence rates are declining in North-America and Europe (Wolters et al., 2020).

Recent developments provide the opportunity for a scoping review to find out more about the body of evidence on the role of social health in the origins of dementia and might provide clues for fine-tuning our hypothesis. Moreover, investigation of the hypothesis requires conceptual clarity. This is lacking for social health where a variety of terms is used interchangeably, namely social network (Berkman, Glass, Brissette, & Seeman, 2000), social integration (Berkman et al., 2000), social engagement (Berkman, 2000), social functioning (Sommerlad, Singleton, Jones, Banerjee, & Livingston, 2017), social capital (Van Deth, 2003) and social contact (Livingston et al., 2020). To make the case for social health as an umbrella concept covering the range of social health characteristics or markers a conceptual framework is required to guide the identification of domains of social health and related markers.

We performed a scoping and a systematic review to underpin our overarching hypothesis and the conceptual framework for social health. We aim to fine-tune our overarching hypothesis on the relationship between social health and cognitive function, to advance concept development through a conceptual framework for social health and to test the initial specified hypothesis: social health affects brain structure and consequently influences cognitive function.

This paper has potential to improve knowledge on the interaction between neurobiological and social health markers and thereby the identification of modifiable risk factors, which may in turn shape new avenues for preventive interventions.

## METHODS

### Study context

The INTERDEM network (pan-European multidisciplinary network of dementia researchers) (www.interdem.org) profiled social health (de Vugt & Droes, 2017; Dröes et al., 2017; M. Vernooij-Dassen & Jeon, 2016; M. Vernooij-Dassen, Moniz-Cook, & Jeon, 2018) and initiated the ‘Social Health And Reserve in the Dementia patient journey (SHARED)’ project, funded by the Joint Programme Neurodegenerative Diseases (JPND) (www.shared-dementia.europe) and its Dutch “sister” project Social Health in Mice and Men (SHiMMy) (Kas, 2018). This paper is part of the SHARED project and uses the first results of the SHiMMy study. The following disciplines are involved in SHARED: epidemiology, sociology, psychology, nursing, geriatrics, psychiatry, neuropsychiatry and family practice. SHARED examines 20 epidemiological studies.

### Design

We used an exploratory design describing the process of hypothesis and concept advancement as follows:

- A scoping and systematic review to underpin and fine-tune our overarching hypothesis and concept advancement
- Concept advancement through the development of a conceptual framework for social health
- Initial testing of a specification of the overarching hypothesis.

The hypothesis and the conceptual framework were iteratively developed by the SHARED team within regular online meetings and a dedicated face-to-face consortium meeting.

#### Scoping review to underpin and fine-tune hypothesis and conceptual framework

The underpinning comprised a scoping review of theoretical and conceptual models, reviews on associations between *single social health markers* and cognitive functioning, and a systematic review on associations between a *combination of social health markers* and cognitive decline or incident dementia.

##### Theoretical and conceptual models

We do not aim to provide a complete overview of models, but to select those which provide insight on the relationship between social health, cognitive decline and onset of dementia.

##### Review on s*ingle* social health markers and cognitive functioning

Studies on associations between *single* social characteristics and cognitive decline and dementia have already been analyzed in reviews. We therefore used reviews to extract information on these associations. However, given that social health is an umbrella concept we want to learn more on the value and power of a *combination* of social health markers. Therefore we additionally performed a new systematic review on the associations of *combinations* of social health markers and cognitive decline and dementia.

##### Systematic review of combinations of social health markers and cognitive decline and dementia

We conducted a systematic database search in the PubMed database from January 2017 through May 2020. The search terms included suitable MeSH terms and keywords on: 1) Cognitive function (e.g., cognitive decline) 2) Social characteristics (e.g., social engagement, 3) Outcome of studies (e.g. associations, longitudinal studies) 4) Study types (e.g. longitudinal). The key inclusion criterion for studies was exploration of the association of a combination of at least two quantitatively measured social health markers with the cognitive functioning outcome (cognitive decline and dementia). Studies not meeting inclusion criteria were excluded. See figure 1 flowchart and appendix on search strategy.

**Figure 1.**
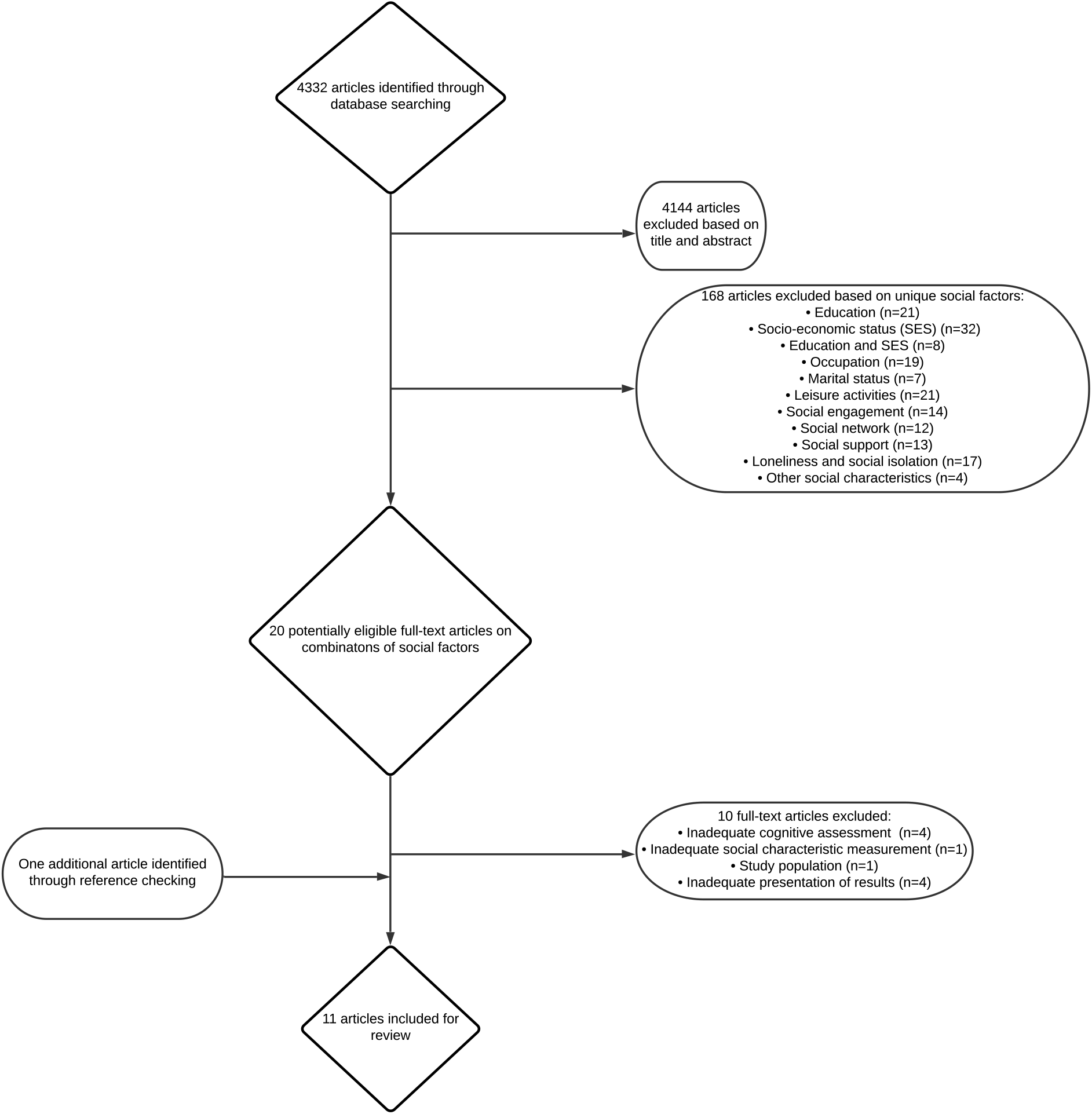
Flowchart

To report the systematic review, we used the PRISMA checklist (Moher, Liberati, Tetzlaff, & Altman, 2009) and for the quality assessment of included studies we used the Newcastle-Ottawa Scale (Wells G, 2009).

##### Concept advancement through the development of a conceptual framework for social health

Concept advancement is a strategic concept –driven effort, that incrementally builds from a conceptual meaning to a more precisely defined unit of meaning (Penrod & Hupcey, 2005). In that way we built a conceptual framework as a system to organize thinking on this complex phenomenon (Emans, 1970-71). We streamlined the route from the core definition towards its constituent domains at the levels of the individual and the social environment, allowing the identification of specific social health markers.

##### Initial hypothesis testing

The process of identification of specific pathways started with the testing of the specified hypothesis within the SHiMMy study using data from the Rotterdam study (van der Velpen et al., 2021). For the social health marker ‘loneliness’ we used a single-item question from the Center for Epidemiological Studies Depression scale and for perceived social support we used a 5-item questionnaire modified from the Health and Lifestyle Survey and marital status. Brain structure was measured by magnetic resonance imaging of the brain to obtain volumetrics, cerebral small vessels disease markers and white matter microstructural integrity (van der Velpen et al., 2021).

## Results

### Underpinning and fine-tuning of hypothesis

We selected theoretical and conceptual work that provide insight in the relationship between social health, cognitive decline and onset of dementia. Thereby we excluded those who did not refer to the role of social health in cognitive decline and the onset of dementia, such as the vascular hypothesis (Penninkilampi, Casey, Singh, & Brodaty, 2018) and a theory relating stress to dementia (Sindi et al., 2017). Most intriguing and relevant to our aim is the cognitive reserve hypothesis (L. Fratiglioni & H.-X. Wang, 2007; Richards & Deary, 2005; Stern et al., 2020). Cognitive reserve reflects the adaptability of the brain that helps to maintain cognitive abilities or day-to-day function despite brain aging, pathology, or injury (Stern et al., 2020; Stern, Barnes, Grady, Jones, & Raz, 2019). It suggests that the brain actively attempts to cope with pathology by using preexisting cognitive-processing approaches or compensatory mechanisms (Stern, 2012).

Fundamentally the cognitive reserve concept is one of effect modification: for a given level of neuropathology, this is less clinically expressed (including, but not limited to, cognitive function) in those with higher ‘reserve’ where education is the original marker) (Richards & Deary, 2005).

But what drives cognitive reserve? Education (L. Fratiglioni & H.-X. Wang, 2007) and more recently social contact (Livingston et al., 2020), an important social health marker, are emerging as factors associated with cognitive reserve.

Our previous hypothesis connecting social health with cognitive decline and dementia departed from the social health conceptualisation of Huber et al. with a strong emphasis on the active role of the individual (Huber et al., 2011; M. Vernooij-Dassen et al., 2021). The importance of the social structure was emphasized in the social network model. The social network model rests on the assumption that the social structure of the network itself shapes the flow of resources which determines access to opportunities and constraints on behaviour (Berkman et al., 2000). The model indicates pathways by which networks might influence health status, but does not include cognitive health status (Berkman et al., 2000).

<u>Reviews of epidemiological studies on s*ingle*</u> social health markers and cognitive functioning demonstrated a link of single social health markers to cognitive decline and dementia (Bellou et al., 2017; Fratiglioni, Paillard-Borg, & Winblad, 2004; L. Fratiglioni & H. X. Wang, 2007; Kelly et al., 2017; Kuiper et al., 2015; Livingston et al., 2020; Livingston et al., 2017; Penninkilampi et al., 2018; Rafnsson, Orrell, d’Orsi, Hogervorst, & Steptoe, 2020; Sutin, Stephan, Luchetti, & Terracciano, 2018). Social health markers including frequency of contacts, social engagement, social network and leisure activities were associated to better cognitive function and may contribute to cognitive reserve and maintenance of cognitive function in old age function (Kelly et al., 2017; Livingston et al., 2020; Penninkilampi et al., 2018) and delay of dementia-symptoms (Fratiglioni et al., 2004; Livingston et al., 2020; Livingston et al., 2017). Conversely, markers of poor social health, such as poor social engagement and social isolation, have been associated with incident dementia (Kuiper et al., 2015; Livingston et al., 2020; Penninkilampi et al., 2018; Piolatto et al., 2022; Rafnsson et al., 2020; Sutin et al., 2018). Recent prospective epidemiological research found that the number of close contacts and being married were associated with a reduced dementia risk (Rafnsson et al., 2020). The results of the influence of loneliness were inconsistent with some reviews concluding that there was no influence (Livingston et al., 2020; Penninkilampi et al., 2018) and recent prospective studies indicating that loneliness was associated with an increased dementia risk (Rafnsson et al., 2020; Sutin et al., 2018). Social isolation has been indicated as one of the 12 risk factors that might prevent or delay the onset in up to 40% of all dementias (Livingston et al., 2020).

<u>The systematic review of combination of social health markers</u> and cognitive decline and dementia yielded 4332 potentially relevant citations, due to the broad search strategy. Based on title and abstract 4144 articles were excluded and 168 potentially eligible full text articles were selected. After excluding articles that did not meet the inclusion criteria, eleven articles were eligible. See figure 1 flowchart. The results indicated that a combination of markers (including ‘social isolation’ and ‘living alone’) was significantly associated with both the development of dementia and reduced cognitive functioning (Ma, Sun, & Tang, 2018; Tsutsumimoto et al., 2017; Yang, Yeung, & Feng, 2018). Combinations including having high occupational complexity and participation in leisure activities turned out to be protective for cognitive functioning (Fujishiro et al., 2019; Opdebeeck et al., 2018; Zhu, Qiu, Zeng, & Li, 2017). This protective function was also present for the development of dementia (Almeida-Meza, Steptoe, & Cadar, 2021; Wang, MacDonald, Dekhtyar, & Fratiglioni, 2017). Engagement in mental activities moderated the impact of widowhood on cognitive functioning (Lee, Chi, & L, 2019). Saito et al. found that 5 social relationship domains were related to a lower likelihood of developing incident dementia: being married, exchanging support with family members, having contact with friends, participating in community groups and engaging in paid work. Combined scores, ranging from 0 to 5, were linearly associated with incident dementia. Those who scored highest were 46% less likely to develop incident dementia compared with those in the lowest category (Saito, Murata, Saito, Takeda, & Kondo, 2018). The effect of using a cumulative score was also highlighted by Tomata et al. who found no significant association with increased dementia risk for low education and living alone separately, but combined with seven other risk factors they demonstrated a dose-response relationship (Tomata et al., 2020). A combination of markers, including social health and other markers, might to be more powerful and influential than single social health markers (Hubers, 2020).

The quality assessment using the Newcastle Ottawa Scale indicated that most methodological flaws were found on the quality items for ascertainment of exposure because of using written self-report (3 of 11 studies) and demonstration that outcome of interest was not present at the start of the study (3 of 11 studies) (Hubers, 2020).

Based on the results of the reviews we explicitly included the role of both the individual and the social environment in the hypothesis:

*Social health is a driver for stimulating the use of cognitive reserve through active facilitation and utilization of the individual’s capacities and those of their social environment that slows cognitive impairment or maintains cognitive functioning in old age*.

A potential working mechanism suggested by Berkman is that social engagement is likely to challenge people to communicate effectively and to engender the mobilisation of cognitive capacities setting in place a ‘use it or lose it’ principle (Berkman, 2000).

The overarching hypothesis is the foundation for formulating verifiable hypotheses to identify modifiable risk factors that can be addressed in future interventions.

### Concept advancement through the development of a conceptual framework for social health

Based on the results of the scoping review we built a conceptual framework including the capacities of the individual and the social environment in the conceptual meaning. We consequently acknowledged the levels of the individual and the social environment and indicated more specific domains on each level. (See figure 2 diagram). This allows to the identification of specific social health markers.

**Figure 2.**
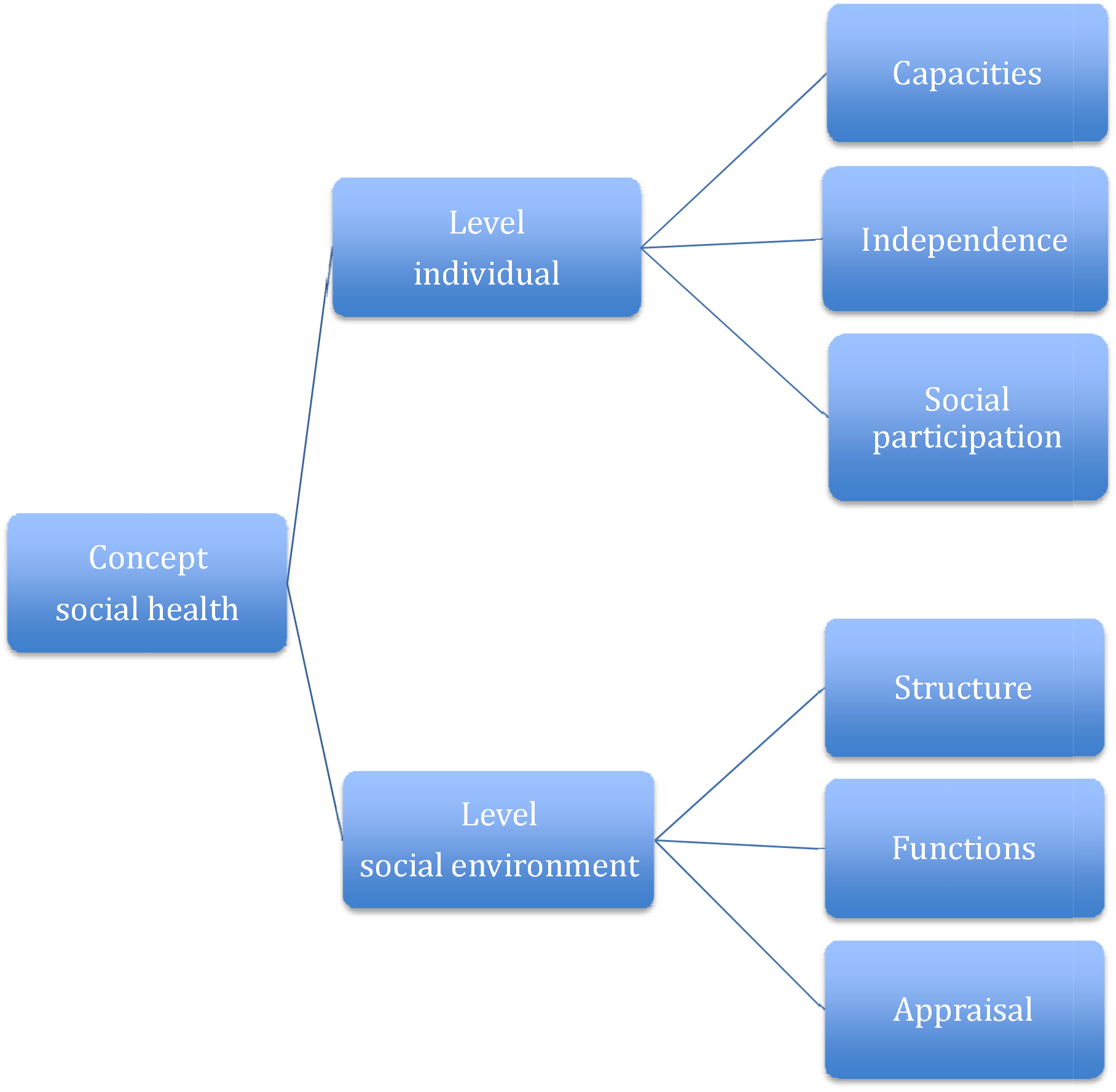
Diagram

The results of the scoping review were consistent with the key sociological referents of relevant *social* phenomena being the consequences of human interactions and the influence of one individual on another (de Jager H, 1974). Social health is not a new term but was defined in 1946 as the social part of the health definition: ‘a state of complete physical, mental and *social well-being*’ (WHO, 1946).

We define social health as *a reciprocal relational concept in which well-being is defined by how an individual relates to their social environment and how the social environment relates to the individual*. This means that social health is essentially a relational concept that refers, on one hand, to the impact that an individual has on others (social environment), and to the impact that the social environment has, in turn, on the individual.

*The individual level*: this represents the competences of the individual to act in social life. Huber et al. identified three domains (Huber et al., 2011): *the capacity to fulfil one’s potential* (according to one’s abilities and talents) *and obligations* (i.e. social demands). This refers to compliance with social norms (Dröes et al., 2017); *the ability to manage life with some degree of independence* (i.e. preserving autonomy and solving problems in daily life). Autonomy refers to acting according to one’s own free will, following own norms and habits. Loss of autonomy can be recognized by dependency, losing control and limitation of activities (M. J. Vernooij-Dassen, Osse, Schade, & Grol, 2005).

The *ability to actively participate in social activities* refers to joining activities with other persons and is underpinned by both theory and epidemiological evidence (L. Fratiglioni & H.-X. Wang, 2007; Livingston et al., 2020; Marseglia et al., 2020).

*The social environment level:* we define its domains in terms of structure, function and appraisal of the quality of the relationship and interaction. Examples of *structural* markers are family or friends’ networks and network diversity (Antonucci, Ajrouch, & Webster, 2019; Ellwardt, van Tilburg, Aartsen, Wittek, & Steverink, 2015). There might be a relevance hierarchy in *structural* markers. It has been suggested that the size of the social network may be less important than its availability (Kuiper et al., 2015). Having varied sources of network ties (such as family and friends) has been found to be beneficial to the cognitive health of older adults over time, while those in networks based mainly on ties with relatives have an increased risk for cognitive decline (Cohn-Schwartz, Levinsky, & Litwin, 2020). Social networks serve a variety of *functions*, including emotional support and instrumental aid (Berkman et al., 2000; Pescosolido) of which the associations with cognitive functioning have been demonstrated (Kelly et al., 2017). *Appraisal* of the quality of the relationship and interaction. Examples of its markers are relationship satisfaction, loneliness, social relationship strain, relationship quality and satisfaction. The appraisal might be positive or negative. Social contacts can be challenging (Walen HR, 2000).

With this conceptual framework we emphasize that an individual’s functioning depends not only on the individual’s own capacities, but also on the social environment’s supportive or hindering behavior. This is crucial in cognitive decline. When sharing these insights at a Patient Public Involvement (PPI) meeting, people with cognitive decline and perceived this information as recognition of their situation: ‘*It is not only about what we cannot do, but also about how others prevent us doing things we can do*’.

### Initial testing of hypothesis

In this initial testing of the overarching hypothesis the social health marker used is loneliness, representing the appraisal of relationships. The hypothesis: social health affects brain structure and consequently influences cognitive function has been tested using the first results of the SHiMMY study based on data from the Rotterdam study (*n* = 3917).

We found that loneliness was associated with smaller white matter volume at baseline (mean difference -4.63 mL, 95%CI: -8.46; -0.81] (van der Velpen et al., 2021). Better perceived social support was associated with larger total brain volume and grey matter volume at baseline and a less steep decrease in total brain volume over time. Better social support was also associated with higher global fractional anisotropy and lower mean diffusivity at baseline. Participants who had never been married had smaller total brain volume (mean difference -8.27 mL, 95%CI -13.16; -3.39) at baseline compared to married peers. In conclusion, social health was associated with brain structure where better perceived social support at baseline was associated with better brain structure over time (van der Velpen et al., 2021).

## Discussion

Our hypothesis linking neurobiological and social sciences, extends thinking beyond a predominantly biological concept and touches upon the complexity of dementia etiology.

While we departed from the social health conceptualisation with a strong emphasis on the role of the individual (Huber et al., 2011; M. Vernooij-Dassen et al., 2021), the importance of the social structure was emphasized in the social network model (Berkman et al., 2000). The reviews on single social health markers revealed associations of individual and social environment markers with maintenance of cognitive function in old age (Livingston et al., 2020) and the delay of dementia-symptoms (Fratiglioni et al., 2004), (Livingston et al., 2017) and cconversely of markers of poor social health with incident dementia (Kuiper et al., 2015; Penninkilampi et al., 2018; Rafnsson et al., 2020; Sutin et al., 2018). The relevance of using an umbrella concept was supported by findings that a combination of markers might to be more powerful in influencing cognitive decline and dementia than single social health markers (Hubers, 2020).

Thus the scoping review suggested the inclusion of both the capacities the individual and the social environment in the hypothesis and the conceptual model for social health and encourages further research on the role of social health in cognitive decline and dementia. In our hypothesis *social health is a driver for stimulating the use of cognitive reserve through active facilitation and utilization of the individual’s capacities and those of their social environment*. Our conceptual framework for social health profiles social health as a relational concept with both the individual and social environment as active parties. The initial testing of our specified hypothesis revealed that better perceived social support at baseline was associated with better brain structure over time (van der Velpen et al., 2021).

The feasibility of the overarching hypothesis was demonstrated by its translation into a specified one. The feasibility of the conceptual framework was demonstrated by its guidance in the identification of social health markers in the SHARED project. This includes the study on mapping of factors influencing cognitive function at the onset of dementia (Seifert I, 2022), an ongoing review on the identification of social health measures, epidemiological studies on associations between social health markers and cognitive decline and dementia and qualitative studies to probe additional relevant social factors. The feasibility of the hypothesis and conceptual framework in the SHJARED project indicates its potential for other social health related dementia studies.

The conceptual framework remains flexible for the novel ideas that can fit under the umbrella concept. Thus, there is scope for the organic growth of new knowledge in this area.

This exploration adds that social health is constituted by both the capacities of the individual and the social environment. Our work expands the 2020 Lancet Commission Report (Livingston et al., 2020) by providing an overarching hypothesis and a conceptual framework to identify social health markers and thereby potential social risk factors (Livingston et al., 2020).

The relevance of social health has actually been demonstrated during The COVID -19 pandemic, which arguably is a large *defacto* natural experiment of social isolation imposed by social distancing and quarantine (M. Vernooij-Dassen, Verhey, & Lapid, 2020). Social isolation has indeed been shown to have a negative impact on cognitive functioning (Almeria, Cejudo, Sotoca, Deus, & Krupinski, 2020; Rohr et al., 2020).

### Delineations

This explorative study has its delineations. The conceptual framework is not a model indicating pathways to social health, but a description of what social health is. For instance, personal characteristics such as introversion and extraversion are not considered to be constituent parts but factors influencing social health. In addition, we see social health from the perspective of the individual and their immediate circle. With this focus, we do not include the larger society and its socioeconomic and cultural characteristics. These are considered to be factors influencing social health (Majoka & Schimming, 2021). An important delineation in this paper is the focus on the onset dementia. We do not miss the perspective of the important role of social health in living with dementia (Samtani, Stevens, & Brodaty, 2021).

### Limitations

There is a diversity of limitations.

The studies included in the reviews use associations and thus we cannot rule out reverse causality.

This conceptual framework is a typical western one, which is illustrated by its emphasis on the value of autonomy. A more diverse perspective should be applied in future work (van Wezel et al., 2016). Moreover, in this conceptualization we did not include the personal needs for social interaction. It is not simply a case of “one size fits all’. Personal characteristics and needs might influence the desire for social contacts and introvert people might enjoy fewer social contacts than others. Intervention studies should adopt an individually tailored approach (Lisko et al., 2020).

A significant limitation regarding the testing of the hypothesis is that there is a gap between the concept of cognitive reserve and its biological substrate and measurement (Richards & Deary, 2005). This limitation has been demonstrated in the testing of our hypothesis in the Rotterdam study where the effect of social health on the relation between MRI-assessed brain pathology and cognitive functioning was not examined (van der Velpen et al., 2021).

### Interventions

The goal is to address modifiable social health risk factors in preventive interventions. Interventions can take place on a clinical level and at a public health level by raising awareness on the importance of social participation for brain health and addressing barriers to social interactions. Children could be taught to develop networks as resources for later life.

The results of the first large dementia prevention trial, the Finnish Geriatric Intervention Study to Prevent Cognitive Impairment and Disability (FINGER) indicated that multi-domain lifestyle-based intervention including stimulation of social interaction by activities could prevent cognitive and functional decline amongst at-risk older adults (Kivipelto et al., 2013; Ngandu et al., 2015).

### Conclusion

The value of social interaction should not be underestimated. Although individual preferences in needs for contact should be considered, social isolation should be prevented. Social contacts are for people, like water to plants. Its value goes beyond “gezelligheid”, the untranslatable Dutch word for enjoying being together, which is seen as an incentive to the ‘thinking activities’ required during social encounters.

## Supporting information

Appendix Search strategy

## Data Availability

All data produced in the present study are available upon reasonable request to the authors

## Acknowledgment

the paper is part of the SHARED project funded by JPND 733051082

## References

Almeida-Meza, P., Steptoe, A., & Cadar, D. (2021). Markers of cognitive reserve and dementia incidence in the English Longitudinal Study of Ageing. Br J Psychiatry, 218(5), 243–251. doi:10.1192/bjp.2020.54

Almeria, M., Cejudo, J. C., Sotoca, J., Deus, J., & Krupinski, J. (2020). Cognitive profile following COVID-19 infection: Clinical predictors leading to neuropsychological impairment. Brain Behav Immun Health, 9, 100163. doi:10.1016/j.bbih.2020.100163

Antonucci, T. C., Ajrouch, K. J., & Webster, N. J. (2019). Convoys of social relations: Cohort similarities and differences over 25 years. Psychol Aging, 34(8), 1158–1169. doi:10.1037/pag0000375

Bellou, V., Belbasis, L., Tzoulaki, I., Middleton, L. T., Ioannidis, J. P. A., & Evangelou, E. (2017). Systematic evaluation of the associations between environmental risk factors and dementia: An umbrella review of systematic reviews and meta-analyses. Alzheimers Dement, 13(4), 406–418. doi:10.1016/j.jalz.2016.07.152

Berkman, L. F. (2000). Which influences cognitive function: living alone or being alone? Lancet, 355(9212), 1291–1292. doi:10.1016/s0140-6736(00)02107-3

Berkman, L. F., Glass, T., Brissette, I., & Seeman, T. E. (2000). From social integration to health: Durkheim in the new millennium. Soc Sci Med, 51(6), 843–857. doi:10.1016/s0277-9536(00)00065-4

Cohn-Schwartz, E., Levinsky, M., & Litwin, H. (2020). Social network type and subsequent cognitive health among older Europeans. Int Psychogeriatr, 1–10. doi:10.1017/s1041610220003439

de Jager H, M. A. L. (1974). Grondbeginselen der sociologie (Basic Principles of sociology). Leiden.

de Vugt, M., & Droes, R. M. (2017). Social health in dementia. Towards a positive dementia discourse. Aging Ment Health, 21(1), 1–3. doi:10.1080/13607863.2016.1262822

Dröes, R., Chattat, R., Diaz, A., Gove, D., Graff, M., Murphy, K., … Johannessen, A. (2017). Social health and dementia: a European consensus on the operationalization of the concept and directions for research and practice. Aging & mental health, 21(1), 4–17.

Ellwardt, L., van Tilburg, T., Aartsen, M., Wittek, R., & Steverink, N. (2015). Personal networks and mortality risk in older adults: a twenty-year longitudinal study. PLoS One, 10(3), e0116731. doi:10.1371/journal.pone.0116731

Emans, R. (1970-71). A schema for classifation of conceptual frameworks involving reading Journal of reading behavior, 3, 15–21.

Fratiglioni, L., Paillard-Borg, S., & Winblad, B. (2004). An active and socially integrated lifestyle in late life might protect against dementia. Lancet Neurol, 3(6), 343–353. doi:10.1016/s1474-4422(04)00767-7

Fratiglioni, L., & Wang, H.-X. (2007). Brain reserve hypothesis in dementia. Journal of Alzheimer’s disease, 12(1), 11–22.

Fratiglioni, L., & Wang, H. X. (2007). Brain reserve hypothesis in dementia. J Alzheimers Dis, 12(1), 11–22. doi:10.3233/jad-2007-12103

Fujishiro, K., MacDonald, L. A., Crowe, M., McClure, L. A., Howard, V. J., & Wadley, V. G. (2019). The Role of Occupation in Explaining Cognitive Functioning in Later Life: Education and Occupational Complexity in a U.S. National Sample of Black and White Men and Women. J Gerontol B Psychol Sci Soc Sci, 74(7), 1189–1199. doi:10.1093/geronb/gbx112

Hardy, J., & Selkoe, D. J. (2002). The amyloid hypothesis of Alzheimer’s disease: progress and problems on the road to therapeutics. Science, 297(5580), 353–356. doi:10.1126/science.1072994

Huber, M., Knottnerus, J. A., Green, L., van der Horst, H., Jadad, A. R., Kromhout, D., … van der Meer, J. W. (2011). How should we define health? BMJ: British Medical Journal, 343.

Hubers, C. (2020). Social characteristics and cognitive function: a systematic review.

Kas, M., Eisel, U., Ikram, A., Vernooij, M., Melis, R., Perry, M., Vernooij-Dassen, M.. (2018). Social Health in Men and Mice (SHiMMy). In. the Netherlands: Groningen, Rotterdam, Nijmegen: ZonMW memorable program (grant number: 733050831).

Kelly, M. E., Duff, H., Kelly, S., McHugh Power, J. E., Brennan, S., Lawlor, B. A., & Loughrey, D. G. (2017). The impact of social activities, social networks, social support and social relationships on the cognitive functioning of healthy older adults: a systematic review. Syst Rev, 6(1), 259. doi:10.1186/s13643-017-0632-2

Kivipelto, M., Solomon, A., Ahtiluoto, S., Ngandu, T., Lehtisalo, J., Antikainen, R., … Soininen, H. (2013). The Finnish Geriatric Intervention Study to Prevent Cognitive Impairment and Disability (FINGER): study design and progress. Alzheimers Dement, 9(6), 657–665. doi:10.1016/j.jalz.2012.09.012

Kuiper, J. S., Zuidersma, M., Oude Voshaar, R. C., Zuidema, S. U., van den Heuvel, E. R., Stolk, R. P., & Smidt, N. (2015). Social relationships and risk of dementia: A systematic review and meta-analysis of longitudinal cohort studies. Ageing Res Rev, 22, 39–57. doi:10.1016/j.arr.2015.04.006

Lee, Y., Chi, I., & L, A. P. (2019). Widowhood, leisure activity engagement, and cognitive function among older adults. Aging Ment Health, 23(6), 771–780. doi:10.1080/13607863.2018.1450837

Lisko, I., Kulmala, J., Annetorp, M., Ngandu, T., Mangialasche, F., & Kivipelto, M. (2020). How can dementia and disability be prevented in older adults: where are we today and where are we going? J Intern Med. doi:10.1111/joim.13227

Livingston, G., Huntley, J., Sommerlad, A., Ames, D., Ballard, C., Banerjee, S., … Mukadam, N. (2020). Dementia prevention, intervention, and care: 2020 report of the Lancet Commission. Lancet, 396(10248), 413–446. doi:10.1016/s0140-6736(20)30367-6

Livingston, G., Sommerlad, A., Orgeta, V., Costafreda, S. G., Huntley, J., Ames, D., … Mukadam, N. (2017). Dementia prevention, intervention, and care. Lancet, 390(10113), 2673–2734. doi:10.1016/s0140-6736(17)31363-6

Ma, L., Sun, F., & Tang, Z. (2018). Social Frailty Is Associated with Physical Functioning, Cognition, and Depression, and Predicts Mortality. J Nutr Health Aging, 22(8), 989–995. doi:10.1007/s12603-018-1054-0

Majoka, M. A., & Schimming, C. (2021). Effect of Social Determinants of Health on Cognition and Risk of Alzheimer Disease and Related Dementias. Clin Ther. doi:10.1016/j.clinthera.2021.05.005

Marseglia, A., Darin-Mattsson, A., Kalpouzos, G., Grande, G., Fratiglioni, L., Dekhtyar, S., & Xu, W. (2020). Can active life mitigate the impact of diabetes on dementia and brain aging? Alzheimers Dement, 16(11), 1534–1543. doi:10.1002/alz.12142

Moher, D., Liberati, A., Tetzlaff, J., & Altman, D. G. (2009). Preferred reporting items for systematic reviews and meta-analyses: the PRISMA statement. Bmj, 339, b2535. doi:10.1136/bmj.b2535

Ngandu, T., Lehtisalo, J., Solomon, A., Levälahti, E., Ahtiluoto, S., Antikainen, R., … Kivipelto, M. (2015). A 2 year multidomain intervention of diet, exercise, cognitive training, and vascular risk monitoring versus control to prevent cognitive decline in at-risk elderly people (FINGER): a randomised controlled trial. Lancet, 385(9984), 2255–2263. doi:10.1016/s0140-6736(15)60461-5

Opdebeeck, C., Matthews, F. E., Wu, Y. T., Woods, R. T., Brayne, C., & Clare, L. (2018). Cognitive reserve as a moderator of the negative association between mood and cognition: evidence from a population-representative cohort. Psychol Med, 48(1), 61–71. doi:10.1017/s003329171700126x

Penninkilampi, R., Casey, A. N., Singh, M. F., & Brodaty, H. (2018). The Association between Social Engagement, Loneliness, and Risk of Dementia: A Systematic Review and Meta-Analysis. J Alzheimers Dis, 66(4), 1619–1633. doi:10.3233/jad-180439

Penrod, J., & Hupcey, J. E. (2005). Concept advancement: Extending science through concept-driven research. Res Theory Nurs Pract, 19(3), 231–241. doi:10.1891/rtnp.2005.19.3.231

Pescosolido, B. The sociology of social networks (Vol. 20): Sage Publications.

Piolatto, M., Bianchi, F., Rota, M., Marengoni, A., Akbaritabar, A., & Squazzoni, F. (2022). The effect of social relationships on cognitive decline in older adults: an updated systematic review and meta-analysis of longitudinal cohort studies. BMC Public Health, 22(1), 278. doi:10.1186/s12889-022-12567-5

Rafnsson, S. B., Orrell, M., d’Orsi, E., Hogervorst, E., & Steptoe, A. (2020). Loneliness, Social Integration, and Incident Dementia Over 6 Years: Prospective Findings From the English Longitudinal Study of Ageing. J Gerontol B Psychol Sci Soc Sci, 75(1), 114–124. doi:10.1093/geronb/gbx087

Richard, E., den Brok, M., & van Gool, W. A. (2021). Bayes analysis supports null hypothesis of anti-amyloid beta therapy in Alzheimer’s disease. Alzheimers Dement, 17(6), 1051–1055. doi:10.1002/alz.12379

Richards, M., & Deary, I. J. (2005). A life course approach to cognitive reserve: a model for cognitive aging and development? Ann Neurol, 58(4), 617–622. doi:10.1002/ana.20637

Rohr, S., Muller, F., Jung, F., Apfelbacher, C., Seidler, A., & Riedel-Heller, S. G. (2020). [Psychosocial Impact of Quarantine Measures During Serious Coronavirus Outbreaks: A Rapid Review]. Psychiatr Prax, 47(4), 179–189. doi:10.1055/a-1159-5562

Saito, T., Murata, C., Saito, M., Takeda, T., & Kondo, K. (2018). Influence of social relationship domains and their combinations on incident dementia: a prospective cohort study. J Epidemiol Community Health, 72(1), 7–12. doi:10.1136/jech-2017-209811

Samtani, S., Stevens, A., & Brodaty, H. (2021). Preserving and enhancing social health in neurocognitive disorders. Curr Opin Psychiatry, 34(2), 157–164. doi:10.1097/yco.0000000000000683

Seifert I, W. H., Lenart-Bugla M, Luc M, Pawlowski M, Rouwette E, Rymaszewska J, Vernooij-Dassen M, Perry M, Melis R, Wolf-Ostermann,Gerhardus A. (2022). Mapping the complexity of dementia: Factors influencing cognitive function at the onset of dementia. BMC Geriatrics, Accepted for publication.

Sindi, S., Kåreholt, I., Solomon, A., Hooshmand, B., Soininen, H., & Kivipelto, M. (2017). Midlife work-related stress is associated with late-life cognition. J Neurol, 264(9), 1996–2002. doi:10.1007/s00415-017-8571-3

Sommerlad, A., Singleton, D., Jones, R., Banerjee, S., & Livingston, G. (2017). Development of an instrument to assess social functioning in dementia: The Social Functioning in Dementia scale (SF-DEM). Alzheimers Dement (Amst), 7, 88–98. doi:10.1016/j.dadm.2017.02.001

Stern, Y. (2012). Cognitive reserve in ageing and Alzheimer’s disease. Lancet Neurol, 11(11), 1006–1012. doi:10.1016/s1474-4422(12)70191-6

Stern, Y., Arenaza-Urquijo, E. M., Bartrés-Faz, D., Belleville, S., Cantilon, M., Chetelat, G., … Vuoksimaa, E. (2020). Whitepaper: Defining and investigating cognitive reserve, brain reserve, and brain maintenance. Alzheimers Dement, 16(9), 1305–1311. doi:10.1016/j.jalz.2018.07.219

Stern, Y., Barnes, C. A., Grady, C., Jones, R. N., & Raz, N. (2019). Brain reserve, cognitive reserve, compensation, and maintenance: operationalization, validity, and mechanisms of cognitive resilience. Neurobiol Aging, 83, 124–129. doi:10.1016/j.neurobiolaging.2019.03.022

Sutin, A. R., Stephan, Y., Luchetti, M., & Terracciano, A. (2018). Loneliness and Risk of Dementia. J Gerontol B Psychol Sci Soc Sci. doi:10.1093/geronb/gby112

Tomata, Y., Li, X., Karlsson, I. K., Mosing, M. A., Pedersen, N. L., & Hägg, S. (2020). Joint impact of common risk factors on incident dementia: A cohort study of the Swedish Twin Registry. J Intern Med, 288(2), 234–247. doi:10.1111/joim.13071

Tsutsumimoto, K., Doi, T., Makizako, H., Hotta, R., Nakakubo, S., Makino, K., … Shimada, H. (2017). Association of Social Frailty With Both Cognitive and Physical Deficits Among Older People. J Am Med Dir Assoc, 18(7), 603–607. doi:10.1016/j.jamda.2017.02.004

van der Velpen, I. F., Melis, R. J. F., Perry, M., Vernooij-Dassen, M. J. F., Ikram, M. A., & Vernooij, M. W. (2021). Social health is associated with structural brain changes in older adults: the Rotterdam Study. Biol Psychiatry Cogn Neurosci Neuroimaging. doi:10.1016/j.bpsc.2021.01.009

Van Deth, J. (2003). Measuring social capital: Orthodoxies and continuing controversies,. International Journal of Social Research Methodology, 6(1), 79–92 DOI: 10.1080/13645570305057

van Wezel, N., Francke, A. L., Kayan-Acun, E., Ljm Devillé, W., van Grondelle, N. J., & Blom, M. M. (2016). Family care for immigrants with dementia: The perspectives of female family carers living in The Netherlands. Dementia (London), 15(1), 69–84. doi:10.1177/1471301213517703

Vernooij-Dassen, M., & Jeon, Y. H. (2016). Social health and dementia: the power of human capabilities. Int Psychogeriatr, 28(5), 701–703. doi:10.1017/s1041610216000260

Vernooij-Dassen, M., Moniz-Cook, E., & Jeon, Y. H. (2018). Social health in dementia care: harnessing an applied research agenda. Int Psychogeriatr, 30(6), 775–778. doi:10.1017/s1041610217002769

Vernooij-Dassen, M., Moniz-Cook, E., Verhey, F., Chattat, R., Woods, B., Meiland, F., … de Vugt, M. (2021). Bridging the divide between biomedical and psychosocial approaches in dementia research: the 2019 INTERDEM manifesto. Aging Ment Health, 25(2), 206–212. doi:10.1080/13607863.2019.1693968

Vernooij-Dassen, M., Verhey, F., & Lapid, M. (2020). The risks of social distancing for older adults: a call to balance. Int Psychogeriatr, 1–3. doi:10.1017/S1041610220001350

Vernooij-Dassen, M. J., Osse, B. H., Schade, E., & Grol, R. P. (2005). Patient autonomy problems in palliative care: systematic development and evaluation of a questionnaire. J Pain Symptom Manage, 30(3), 264–270. doi:10.1016/j.jpainsymman.2005.03.010

Walen HR, L. M. (2000). Social support and strain from partner, family, and friends:costs and benefitsfor men and womenin adulthood. J Soc Personal Relationships, 17, 5–30.

Wang, H. X., MacDonald, S. W., Dekhtyar, S., & Fratiglioni, L. (2017). Association of lifelong exposure to cognitive reserve-enhancing factors with dementia risk: A community-based cohort study. PLoS Med, 14(3), e1002251. doi:10.1371/journal.pmed.1002251

Wells G, S. B., O-Connell D, Peterson J, Welch V, Losos M e tal. (2009). The Newcastel-Ottawa Scale (NOS) for assessing the quality of non-randomised studies in meta-analyses. Retrieved fro https://www.ohri.ca/programs/clinical_epidemiology/oxford.asp. https://www.ohri.ca/programs/clinical_epidemiology/oxford.asp

WHO. (1946). Preamble to the constitution of WHO as adopted by the International Health Conference. New York 1946. Retrieved from https://www.who.int/about/governance/constitution

Winblad, B., Amouyel, P., Andrieu, S., Ballard, C., Brayne, C., Brodaty, H., … Zetterberg, H. (2016). Defeating Alzheimer’s disease and other dementias: a priority for European science and society. Lancet Neurol, 15(5), 455–532. doi:10.1016/s1474-4422(16)00062-4

Wolters, F. J., Chibnik, L. B., Waziry, R., Anderson, R., Berr, C., Beiser, A., … Hofman, A. (2020). Twenty-seven-year time trends in dementia incidence in Europe and the United States: The Alzheimer Cohorts Consortium. Neurology, 95(5), e519–e531. doi:10.1212/wnl.0000000000010022

Yang, Y., Yeung, W. J., & Feng, Q. (2018). Social exclusion and cognitive impairment - A triple jeopardy for Chinese rural elderly women. Health Place, 53, 117–127. doi:10.1016/j.healthplace.2018.07.013

Zhu, X., Qiu, C., Zeng, Y., & Li, J. (2017). Leisure activities, education, and cognitive impairment in Chinese older adults: a population-based longitudinal study. Int Psychogeriatr, 29(5), 727–739. doi:10.1017/s1041610216001769

