## Appendix Search strategy for "CONCEPTUAL ADVANCEMENT: SOCIAL HEALTH AS A FACILITATOR IN THE USE OF COGNITIVE RESERVE"

### Supplemental file

#### Search strategy

"Dementia"[Mesh:NoExp] OR "Alzheimer Disease"[Mesh] OR "Cognitive Dysfunction"[Mesh:NoExp] OR
Dementia*[tiab] OR Alzheimer*[tiab] OR Cognitive impairment*[tiab] OR Cognitive capacit*[tiab] OR Cognitive reserve[tiab] OR Cognitive decline[tiab] OR Cognitive function*[tiab] OR Cognitive dysfunction*[tiab]

AND

Reciprocity[tiab] OR Social awareness[tiab] OR Capacit*[tiab] OR "Personal Autonomy"[Mesh] OR Independen*[tiab] OR Dependen*[tiab] OR Autonom*[tiab] OR "Activities of Daily Living"[Mesh] OR "Social Participation"[Mesh] OR "Leisure Activities"[Mesh] OR Activities of daily living[tiab] OR Leisure[tiab] OR Social participation[tiab] OR Social engagement[tiab] OR Social disengagement [tiab] OR Social integration[tiab] OR Social activit*[tiab] OR Social withdrawal[tiab] OR "Independent Living"[Mesh] OR "Marital Status"[Mesh] OR "Socioeconomic Factors"[Mesh:NoExp] OR "Employment"[Mesh] OR "Educational Status"[Mesh] OR "Social Environment"[Mesh] OR Independent living[tiab] OR Marital status[tiab] OR Marriage[tiab] OR Socioeconomic factor*[tiab] OR Socioeconomic status [tiab] OR Income[tiab] OR Education[tiab] OR Social environment[tiab] OR Social network structure[tiab] OR Network composition[tiab] OR Living arrangement*[tiab] OR "Social Support"[Mesh] OR "Loneliness"[Mesh] OR "Personal Satisfaction"[Mesh] OR "Interpersonal Relations"[Mesh:NoExp] OR "Social Isolation"[Mesh] OR social support[tiab] OR loneliness[tiab] OR personal satisfaction[tiab] OR interpersonal relation*[tiab] OR social isolation[tiab] OR intimate relation*[tiab] OR relationship quality[tiab] OR personal network[tiab] OR social network[tiab] OR social influence[tiab] OR friendship[tiab] OR family relation*[tiab] OR social tie*[tiab] OR social relation*[tiab] OR social interaction [tiab] OR support system[tiab]

AND

"Risk"[Mesh:NoExp] OR "Risk Factors"[Mesh] OR "Risk Assessment"[Mesh] OR "Prognosis"[Mesh:NoExp] OR "Protective Factors"[Mesh]
OR
Risk*[tiab] OR Risk factor*[tiab] OR Predictive factor*[tiab] OR Cause[tiab] OR Prognostic factor*[tiab] OR Contributing factor*[tiab] OR Predisposing factor*[tiab] OR Protective factor*[tiab] OR Association*[tiab]

AND

"Cohort Studies"[Mesh] OR "Observational Study" [Publication Type] OR "Case-Control Studies"[Mesh]
OR
Cohort[tiab] OR Longitudinal[tiab] OR Prospective[tiab] OR Retrospective[tiab] OR Follow-up[tiab] OR observational[tiab]
